# Pattern of fibrin stabilizing factor (FXIII) expression in placentas of women suffering with gestational diabetes mellitus in Saint John, New Brunswick

**DOI:** 10.1101/2021.10.08.21264740

**Authors:** Victoria J. Mercer, Kenneth Obenson

**Author notes:** Correspondence concerning this article should be addressed to Victoria Mercer, 100 Tucker Park Rd, Saint John NB E2K 5E2, Canada. Author Note We received no funding for this study.

## Abstract

Gestational diabetes mellitus (GDM) affects 2-5% of all pregnancies and is known to place the fetus at risk for adverse fetal outcomes. Previous studies have reported the increased presence of villous immaturity in the placentas of women with gestational diabetes. Villous immaturity poses a great risk of restricted diffusion capacity across the placenta and may be a marker for some of the placental insufficiency associated with diabetes mellitus. The present study looks at the possibility of using fibrin stabilizing factor, or Factor XIII (FXIII), as a biomarker of villous immaturity. The aim of the present study is to establish a baseline pattern of FXIII expression in placentas of women in Saint John, NB with GDM, without villous immaturity using a scoring formula adapted from surgical pathology. While the small sample size precludes definitive conclusions regarding the expression of FXIII in normal placentas in women with GDM, there appears to a baseline of strong FXIII expression in normal placental tissues from the second and third trimester of pregnancy. Further study using a larger sample size is required to determine if a correlation exists between level of FXIII expression and degree of villous maturity in patients with GDM, which could improve the histologic assessment of placental development.

## Introduction

### Effects of diabetes during pregnancy

Gestational diabetes mellitus (GDM) is defined as hyperglycaemia with blood glucose values above normal but below those diagnostic of diabetes and/or impaired glucose tolerance, occurring during pregnancy [1]. In addition to obesity, poor diet and lack of exercise, risk factors for GDM include older maternal age as well as family history of diabetes, multiparity and previous GDM [2].

Gestational diabetes affects 2-5% of all pregnancies and is known to place the fetus at risk for adverse fetal outcomes as well as a general increased risk of fetal morbidity and mortality [1,3]. Fetal complications of GDM include macrosomia, birth trauma, neonatal hypoglycemia as well as respiratory distress syndrome [1].

The placenta is the interface between the mother and fetus and is essential for transmission of oxygen and nutrients. It is subject to the adverse effects of various maternal conditions. The placenta is the only fetal tissue capable of storing excess glucose, suggesting that maternal hyperglycemia can play a direct role in fetal complications of GDM such as macrosomia [4]. Jarmuzek, Wielgos, Bomba-Opon, et al. propose that maternal hyperglycemia resulting in an increased transplacental glucose gradient can produce an enlarged placenta, precipitating transport of nutrients and accelerated fetal growth [3]. This is merely a single illustration of the influence the placenta and its development have on pregnancy outcome.

Studies report that the typical changes seen in the placenta affected by GDM are generally either structural or functional based on when in the pregnancy the mother develops GDM [4]. The extent of these changes additionally depends on the degree of glycemic control [4]. Maternal hyperglycemia stimulates metabolic changes in the fetus such as increased insulin production, increased metabolism, an increase in overall oxygen demands, and subsequently chronic fetal hypoxia [3]. In pregnancies in which the mother has an antepartum condition such as gestational diabetes that has known associations with villous abnormalities, potential correlations with FXIII expression could possibly establish a biomarker to aid in detection of abnormal placental development. Although the importance of FXIII in maintaining pregnancy in a homozygous FXIII deficiency situation has been demonstrated, there are few studies exploring pregnancy outcomes in patients with reduced FXIII levels. The question remains whether or not FXIII levels are associated with adverse outcomes related to villous abnormalities and placental development. The aim of the present study is to establish a baseline pattern of FXIII expression in placentas of women in Saint John, NB with GDM, without villous immaturity. This study was presented as a poster at the 2021 Society of Obstetrics and Gynaecology of Canada’s Annual Clinical and Scientific Conference Medical Student Program on June 19^th^ 2021.

## Study Objectives

1. Establish a baseline of FXIII expression in placentas of women with gestational diabetes mellitus.
2. Develop a method of scoring FXIII expression.

### Participants

Horizon Health Network’s Research and Ethics Board approved this study in February 2020. The study consists of a retrospective cohort of women who gave birth at the Saint John Regional Hospital between January 2014 and December 2020. The principle investigator (PI) searched the Pathology Department’s database for all placentas from mothers with a clinical history of gestational diabetes received at the Pathology Department between January 2014 and December 2020. This study considered both singleton and twin pregnancies. Twenty-five cases without villous immaturity were selected by the PI to be considered for FXIII immunohistochemistry.

Inclusion criteria was a history of gestational diabetes. Exclusion criteria was any other maternal co-risk factor for villous immaturity such as drug or alcohol use, smoking, chromosomal/genetic anomaly, no antenatal care or TORCH (toxoplasmosis, other, rubella, cytomegalovirus, herpes simplex) infections (infections known to produce congenital defects).

Three random cases were selected (from the 25 cases above) by the PI to have two samples each stained with FXIII antibodies. The co-investigator scoring the samples was blinded to the duplicated slides to serve as a method of internal quality assurance. One sample per case was stained from the remaining 22 cases.

## Methods

### Preparation of Placenta Tissues and Slides

From each of the 25 cases one sample was collected. From three of the 25 case, an additional sample was collected as a measure of internal quality control. The result was 28 slides to be stained for FXIII. The samples were selected based on general quality of the stain. Five fields were randomly selected by the co-investigator from across the placenta disc. Fields were limited to examination of terminal villi and did not include sections taken from near the disk edge or areas of infarction, hence the requirement for initial triages by the PI before the immunohistochemistry was ordered. Immunohistochemistry and application of FXIII antibodies was conducted by Horizon Health Pathology Department.

Immunohistochemistry was performed on tissue samples that were sectioned and prepared per usual procedure for tissue processing, following standard fixation and routine tissue embedding protocols.

Once the samples were stained for FXIII and reviewed by the PI for stain quality and adequacy, a member of the Horizon Health Research team that was not in associated with this study, de-identified all samples. The key to re-identify the samples was saved on an encrypted USB that neither the principle nor co-investigator had access to.

### Scoring of FXIII expression

All samples were viewed with Olympus microscope.

A total of five microscope fields per sample at 20x magnification were randomly selected and scored by the co-investigator based on intensity of the stain and the extent of FXIII expression.

The average intensity and extent of staining of the five fields per case was used in accordance with the scoring method depicted in Table 1 to assign an overall score of FXIII expression per sample. The higher the score, the higher the expression of FXIII.

**Table 1.**
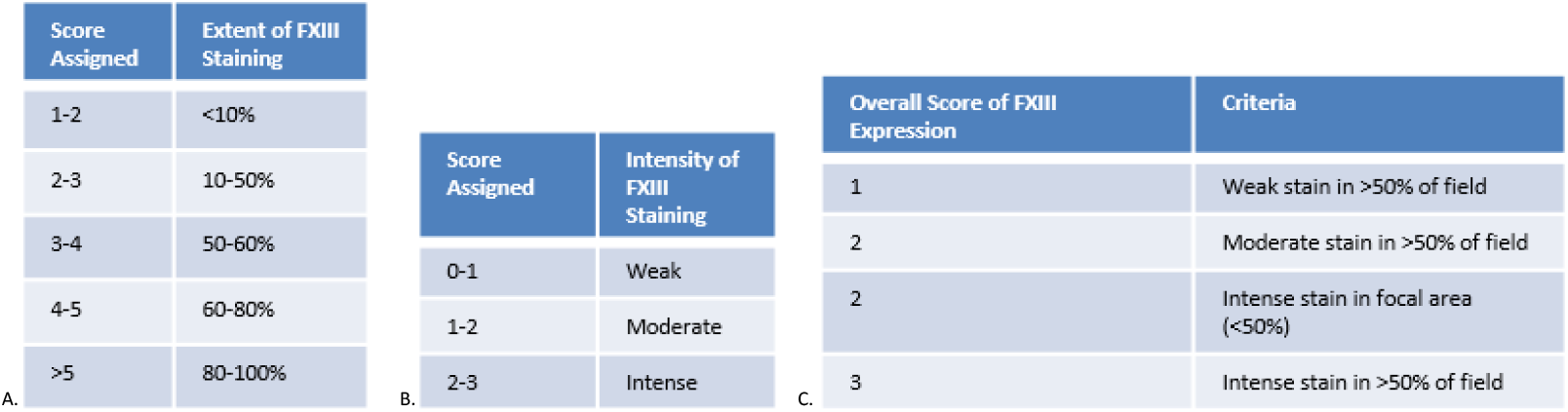
Scoring method of FXIII expression. Using the average extent (a) and intensity (b) of staining. within five separate fields per case allows a representative and objective overall score(c) of FXIII expression to be assigned to each case.

Extent of staining was scored from 1-5 with 1 indicating less than 10% staining of FXIII in a single field, 2 indicating 10-50% staining, 3 indicating 50-60%, 4 indicating 60-80% and a score of 5 indicating >80% of the villi in a single field are stained with FXIII. An average of the extent of expression scores assigned to the five fields per case was considered the overall extent of FXIII expression score for that sample.

Stain intensity was graded in each field from 0-3, 3 being the strongest expression (staining) of FXIII. A score of 0 indicated no staining of FXIII. A score of 1 and 2 indicated little or moderate staining intensity in that field, respectively. An average of the intensity scores assigned to each of the five fields per case was considered the overall intensity score for that sample.

The scores assigned to each field are exclusively whole numbers, however in Table 1 the extent and intensity are depicted as ranges due to the effect of averaging the five scores for each sample. The average intensity and extent of staining for each case was used to assign an overall score of FXIII expression per sample. The higher the score, the higher the expression of FXIII.

All scores were recorded on a Microsoft Excel Sheet (Appendix A) and stored on an encrypted USB drive.

From the 25 cases, three randomly selected cases had two slides each stained with FXIII antibodies as a measure of internal quality assurance, while one slide was stained from the rest of the samples. The co-investigator scoring the FXIII expression was blinded to the duplicate slides.

### Data Collection

During data collection, the information was stored on an encrypted USB. Following data analysis and the completion of this study, all data collected will be transferred to a secured I drive folder in Horizon

Health’s Department of Laboratory Medicine. All information on the encrypted USB will be erased by the co-investigator. No paper documents were required for use during this study.

Standard practice at Horizon Health Network requires the data collected during this non-clinical trial research project to be retained for 7 years. After 7 years all identifying information will be deleted. Any deidentified information may be kept indefinitely.

When all 140 fields are scored (five fields per case, a total of 25 cases plus three duplicated samples also with five fields each), the samples will be re-identified using the key the secretary member of the Pathology Department of the Saint John Regional Hospital created. This will re-associate FXIII expression score with identifying information including adverse pregnancy outcomes.

### Statistics

FXIII extent, intensity and overall scores were averaged to establish a baseline score of FXIII expression in placentas of women with GDM, without villous immaturity.

## Preliminary Results

Out of 25 samples there were no reported cases of villous immaturity. The average strength of FXIII expression was 2.80/3.00.

Nine out of 25 of placentas sampled were ≥90^th^ percentile in placenta weight and of these, seven scored an overall FXIII expression score of three, two scored a two.

Included in the study were two twin pregnancies and 23 singletons. Three of the samples were stained twice as a measure of internal control to ensure consistency and accuracy of the scoring process.

## Discussion

The method of scoring FXIII in this study is an adaptation of the international standard of scoring of hormonal status in breast cancer cases, approved by the Canadian Association of Pathologists. This study aimed to give an objective scheme to an otherwise subjective score of immunohistochemistry staining. The consistency in the overall score of expression of the three controls (Figure 2) within the samples indicates that this scoring method is reliable.

**Figure 1:**
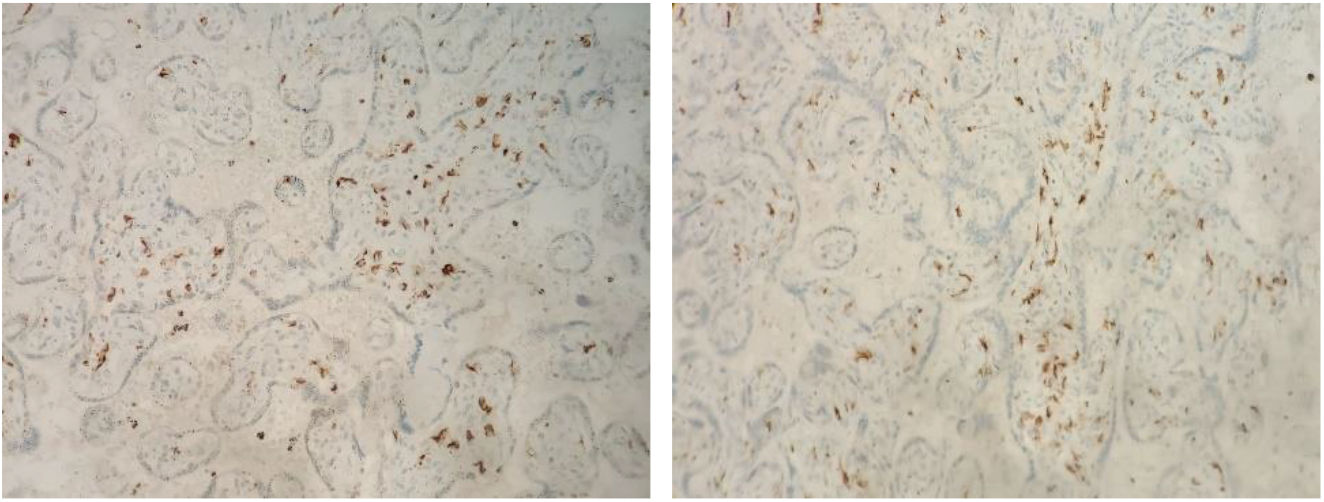
Placenta tissue stained with FXIII viewed at 20x magnification

**Figure 2:**
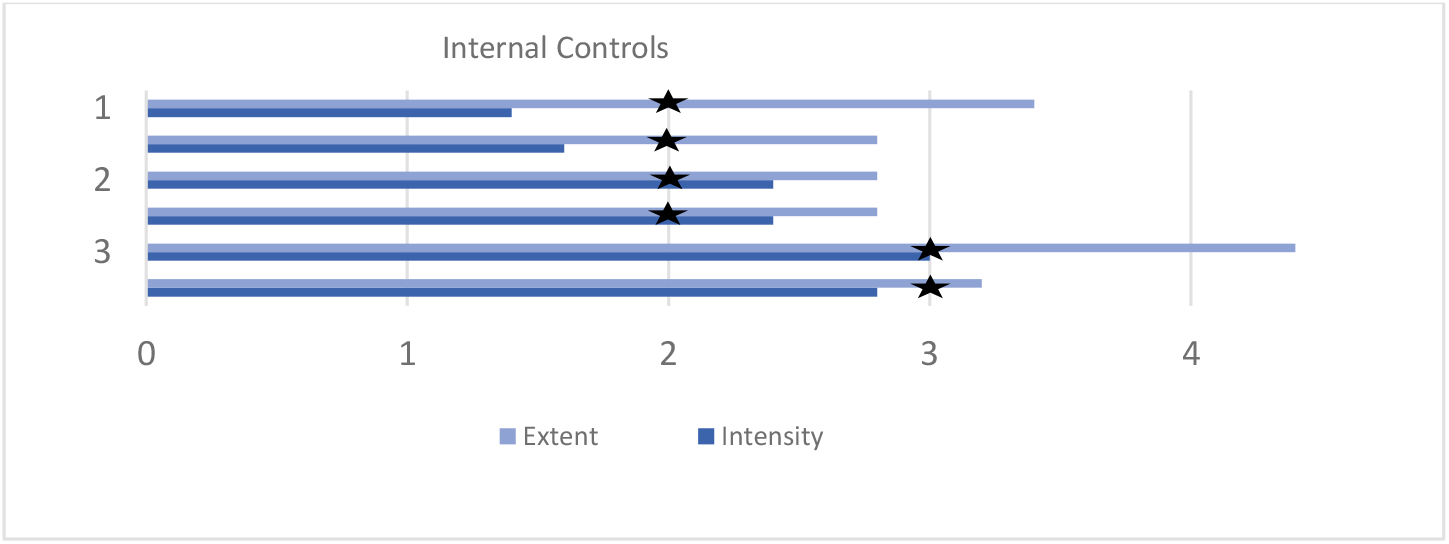
Bar graph representation of averaged FXIII intensity and expression of three controls. Average scores of intensity and extent of each sample vary slightly amongst the randomly selected five fields. Using the average of the five fields of each sample with the criteria described in Table 1, the overall FXIII expression score is identical among each control, indicated by the black star

### Role of FXIII in placental development

Previously known is the necessity for FXIII protein in the maintenance of pregnancy via its role as fibrin stabilizing factor and potentially contributing to angiogenesis. FXIII is a tetrameric protein consisting of two catalytic subunits referred to as FXIIIa, and two carrier subunits (FXIIIb) [5]. FXIII is a zymogen that has a critical role in the regulation of fibrinolysis in addition to facilitating the final stage of the coagulation cascade [6]. FXIIIa forms covalent cross-links between glutamyl and lysyl residue on adjacent fibrin chains, contributing to the strength, rigidity and stiffness of the mature clot, as well as its resistance to fibrinolysis [6]. Most FXIII deficiencies are due to an autosomal recessive mutation in the A-subunit [6]. The characteristic bleeding often occurs from the umbilical cord at birth and intracranially [6].

In addition to an increased occurrence of gross fetus complications, it has been suggested that depending on the degree of glucose tolerance, the microanatomical morphology of the placenta can be affected as well [3,4]. The extensive angiogenesis and vascularization of the placenta that takes place in the second half of pregnancy may be impacted in response to disturbances in glucose metabolism [1,3,4]. Factor XII has been shown to be critical to the development of an adequate vascular bed that will act as a conduit for both nutrients and waste removal. Some of the typical changes in placentas of poorly controlled GDM patients include villous edema, fibrin deposits, hyperplasia in the syncytiotrophoblast as well as general placental immaturity [1,3]. In a study conducted by Jones and Fox, seven placentas from women with gestational diabetes were examined [7]. In five of the seven placentas, the terminal villi were immature for the length of the gestational period (all patients delivered between 38^th^ and 39^th^ week) [7]. In a study conducted by Madazli, Tuten, Calay et al., the presence of villous immaturity, chorangiosis and ischemia were significantly increased in the placentas of women with gestational diabetes [1].

Levels of factors VII, VIII, XI, X, fibrinogen, as well as FXIIIb.tend to increase progressively during pregnancy [5]. During the second half of pregnancy, extensive angiogenesis and vascularization of the placental villi occurs [3]. Dardik, Loscalzo, and Inbal propose that the dimerization of FXIIIa is crucial to forming the cytotrophoblastic shell and Nitabuch’s layer at approximately six to eight weeks gestation [6]. In a study of FXIII levels during pregnancy, it was found that levels were significantly lower in the 31-40^th^ week compared to 0-10 although whether reduction of FXIIIa can be attributed to reduced synthesis/reduced requirement or increased utilization remains unclear [8]. It is clear however that FXIII is essential for maintaining pregnancy. A normal reference range for FXIII is 70-140 IU dL^1^ [8].Women homozygous for FXIII deficiency are unable to carry a pregnancy to term and have been found to suffer from reoccurring miscarriages, generally in the first trimester [5,6].

Latent placental insufficiency, antenatal hypoxia, unexplained intrauterine death and abnormal neonatal outcomes are all common results of idiopathic villous immaturity [9]. Villous immaturity poses a great risk of restricted diffusion capacity, therefore limiting oxygen and nutrient exchange across the placenta. This means that earlier identification of histological changes of the placenta in women with GDM could provide the opportunity for medical intervention and prevent adverse pregnancy outcomes. The present study considers the possibility of using fibrin stabilizing factor, or Factor XIII (FXIII), as a biomarker of villous immaturity by establishing a baseline FXIII expression in placentas of women with GDM.

The established baseline and proven scoring method can be used for future studies to explore possible correlations between FXIII and villous immaturity. Potential implications include using FXIII expression to predict villous immaturity in patients with gestational diabetes and subsequently predict placental competency.

### Study Limitations

A major limitation to the study is the small sample size, which prevented us from reaching any statistically significant conclusions. This limitation presented itself when searching though the Horizon Health Pathology Department’s database for placentas from mothers with a clinical history of GDM who delivered between January 2014 and December 2020. Such cases were rare. In the study conducted by Madazli, Tuten, Calay et al, 22 women with GDM were followed for just over a year in Istanbul, a city with a population size over 15 million - substantially larger than Saint John, NB with a population size of merely 67 500 people [1]. With the incidence of GDM being 2-5%,the small population of Saint John, NB severely limits our access to women with GDM. Furthermore, not every placenta meeting conditions for placenta examination is sent to pathology for a variety of reasons. These may include obstetrician judgement, patient preference, resource management, etc. Collecting more samples over time will provide opportunity to follow up with more cases using this scoring method and to expand this study by examining FXIII expression in abnormal cases, particularly those with villous immaturity.

## Conclusions

Due to the small sample size, conclusions about FXIII expression in normal placenta tissue cannot be made. However preliminary findings have established a baseline of strong FXIII expression in normal placental tissues from the second and third trimester of women with GDM without villous immaturity.

Although definitive conclusions regarding FXIII expression in placentas with or without villous immaturity could not be made, the study has established a reliable method of scoring FXIII expression. Further studies using this methodology with a larger sample size is required to determine if the preliminary baseline of strong FXIII expression in normal placentas will remain consistent. This study provides the introduction and proposed methodology needed to more thoroughly investigate the relationship of FXIII and the placenta.

## Data Availability

All data produced in the present study are available upon request to the authors.

## References

1. Madazli R, Tuten A, Calay Z, Uzun H, Uludag S, Ocak V. The Incidence of Placental Abnormalities, Maternal and Cord Plasma Malondialdehyde and Vascular Endothelial Growth Factor Levels in Women with Gestational Diabetes Mellitus and Nondiabetic Controls. Gynecologic and Obstetric Investigation. 2008;65(4):227–232. 10.1159/000113045

2. Katarzyna Cypryk A, Cypryk K, Szymczak W, Czupryniak L, Sobczak M, Lewiński A. Gestational Diabetes Mellitus-an Analysis of Risk Factors. Polish Journal of Endocrinology. 2008(59):393–397. https://journals.viamedica.pl/endokrynologia_polska/article/view/25528

3. Jarmuzek P, Wielgos M, Bomba-Opon DA, Dorota A, Bomba-Opon A. Placental Pathologic Changes in Gestational Diabetes Mellitus. Neuroendocrinol Lett. 2015;36(2):101–105. https://www.nel.edu/userfiles/articlesnew/NEL360215R01.pdf

4. Desoye G, Hauguel-De Mouzon S. The Human Placenta in Gestational Diabetes Mellitus: The Insulin and Cytokine Network. Diabetes Care. 2008;30(2):120–126. 10.2337/dc07-s203

5. Muszbek L, Yee VC, Hevessy Z. Blood coagulation factor XIII: Structure and function. Thrombosis Research. 1999;94(5):271–305. 10.1016/S0049-3848(99)00023-7

6. Dardik R, Loscalzo J, Inbal A. Factor XIII (FXIII) and angiogenesis. Journal of Thrombosis and Haemostasis. 2006;4(1):19–25. 10.1111/j.1538-7836.2005.01473.x

7. Jones CJP, Fox H. Placental Changes in Gestational Diabetes: An Ultrastructural Study. Obstetrics & Gynecology. 1976;48(3):274–280. https://oce-ovid-com.ezproxy.library.dal.ca/article/00006250-197609000-00004/HTML

8. Sharief LT, Lawrie AS, Mackie IJ, et al. Changes in factor XIII level during pregnancy. Haemophilia. 2014;20:144–148. 10.1111/hae.12345

9. Seidmann L, Suhan T, Kamyshanskiy Y, Nevmerzhitskaya A, Gerein V, Kirkpatrick CJ. CD15 A new marker of pathological villous immaturity of the term placenta. Placenta (Eastbourne). 2014;35(11):925–931. 10.1016/j.placenta.2014.07.018

